# Trends of SARS-CoV-2 infection worldwide: Role of population density, age structure, and climate on transmission and case fatality

**DOI:** 10.1101/2020.05.20.20104257

**Authors:** Tewodros Endailalu, Fitsum Hadgu

## Abstract

**Introduction:** The highly heterogenous disease transmission pattern of COVID-19 suggests that the pandemic maybe driven by complex factors, which may include habitat suitability, region specific human mobility, and transmission related to susceptibility. The purpose of this study was to examine the effects of different spatial and demographic factors on COVID-19 transmission and case fatality worldwide.

**Methods:** We assessed SARS-CoV-2 virus transmission and COVID-19 related fatalities in 50 countries in all continents of the globe. Data from the COVID-19 data repository of the Johns Hopkins Center for Systems Science and Engineering, the European center for disease control and prevention, and the World Health Organization were used to obtain the daily number of cases and organize incidence data. Disease spread was assessed using the reproduction number of the disease across the sampled countries. R statistical software’s R0 package was used to estimate the reproduction number of the COVID19 using the exponential growth method. After computing the reproductive number of each country in the study, a multiple linear regression model was fitted using R0 value as dependent variable, and latitude and population density as an independent variable. Disease severity was analyzed using the case fatality ratio of COVID-19. The proportion of deaths were meta-analyzed using the R statistical software’s metaphor package, using random effect inverse variance weighting to come up with the case fatality ratio.

**Results:** We found no statistically significant association between disease spread and latitude or population density. The regression model analysis that accounted for age, population density and latitude showed that age distribution remains an important driver shaping the current distribution of COVID-19 cases. The relative frequency of people above 65 years old was positively correlated with the cumulative numbers of COVID-19 cases as well as case fatality ratio in each country. The multiple linear regression model fitted between CFR and the three major covariates showed that, the demographic distribution of the sampled countries is strongly associated with the case fatality ratio. Correlation with proportion of populations over 65 years old is concordant with the previous findings relationship between case fatality ratio and patient age.

**Conclusion:** This analysis provides important information that can inform the decisions of local and global health authorities. Particularly, as our study confirms that death and severity of COVID19 are associated with age, in countries with the biggest outbreaks, strategies must be employed to ensure that high-risk groups, such as old people received adequate protection from COVID-19.

## Introduction

The Severe Acute Respiratory Syndrome Coronavirus 2 (termed SARS-CoV-2) pneumonia – also known as Coronavirus Disease 19 (COVID-19) – began to appear in the city of Wuhan, the capital city of the Hubei Province, in Central China (30.60°N – 114.05°E). As of May 15, 2020, at 6:00pm (ET), the Johns Hopkins University Center for Systems Science and Engineering reported that there have been 4,586,915 confirmed cases of COVID-19 infection and 279,597 deaths. In comparison, there were 774 reported deaths from the 2003 SARS outbreak. The five countries hardest hit by COVID-19 have been the United States (1,450,269 cases), Russia (272,043 cases), United Kingdom (241,455 cases), Spain (230,698 cases), and Italy (223,885 cases). [1]

On Jan 30, 2020, WHO declared the current novel coronavirus disease 2019 (COVID-19) epidemic a Public Health Emergency of International Concern. [2] All continents have reported confirmed cases of COVID-19. Africa confirmed its first case in Egypt on Feb 14, 2020. An initial assessment of the outbreak by Li and colleagues estimated that in the early phase of the COVID-19 outbreak in China, the epidemic doubled in size every 7.4 days and the basic reproductive number (R0) was 2.2. [3]

It has been challenging to accurately track the spread, because of factors such as the lack of rapid diagnostic tests and the mildness of the symptoms in some infected people. SARS-CoV-2 shares 88% sequence identity to two coronaviruses found in bats, bat-SLCoVZC45 and bat-SL-CoVZXC21, 79% identity with the Severe Acute Respiratory Syndrome (SARS) coronavirus and 50% identity with Middle Eastern Respiratory Syndrome (MERS) coronavirus.[4] The case fatality ratio (CFR) for cases outside China was initially estimated to be 2.2% (95% confidence interval, 0.6%-5.8%). [5] A recent study of the symptomatic case fatality risk (the probability of dying from the infection after developing symptoms) in Wuhan found that the overall risk was 1.4% in patients aged 15 years or older. [6]

The virus has rapidly spread worldwide, and future scenarios remain highly uncertain. Modelling by the Imperial College COVID-19 Response Team in London predicted approximately 510,000 deaths in Great Britain and 2.2 million in the US, not accounting for the potential negative effects of health systems being overwhelmed on mortality. [7] This is argued as a worst-case scenario, which even if true would be mitigated by the many people who would be minimally or mildly symptomatic.

The disease transmission pattern of COVID-19 is highly heterogeneous suggesting that the pandemic maybe driven by complex factors, which may include habitat suitability, region-specific human mobility, and transmission related to susceptibility.[8] Some countries such as Vietnam had cases from the earliest stage of the outbreak but experienced a relatively moderate increase in new cases, whereas others such as the UK and the USA have been suffering serious epidemic. Therefore, identifying the driving force of the outbreak pattern is urgently needed for predicting infection and mortality risks in the remaining less affected countries of the world.

In terms of mortality, case fatality rate for COVID-19 have been previously shown to be associated with age, comorbidities and sex. [9] Other studies also suggest that country-specific case fatality ration and incidence are corelated with latitude and temperature. [10-11] Several other studies show that population age structure may explain the variation in fatalities across countries. For example, Italy is one of the oldest populations, with 23.3% of its population over 65 years, characterized by significant intergenerational contacts and residential proximity between adult children and parents. [12] The Korean outbreak was also concentrated among the young Shincheonji religious group, with only 4.5% of cases thus far falling into the >80-y group. [13] This contributed to a low overall CFR in South Korea relative to Italy (1.6% vs. 10.6%). The hypothesis that warmer temperatures will slow the spread of the COVID-19 virus has been a point of much debate. However, early comparisons between the number of confirmed cases in the worst affected European countries and the West African countries with confirmed COVID-19 cases do not support the hypothesis that the virus will spread more slowly in countries with warmer climates. [14] The purpose of this study was to examine the effects of different spatial and demographic factors on COVID-19 transmission and case fatality worldwide.

## Methods

We assessed SARS-Cov-2 virus transmission and COVID-19 related fatalities in 50 countries in all continents of the globe.

### Sampling

More than 200 countries and territories that have reported the virus were initially included as “target population”. We first excluded countries that have not reported COVID-19 related death, as of April 29, leaving the eligible sampling frame size at 172. In the same way, to better understand the transmission of the virus and allow of meaningful comparison and interpretation of the results countries that reported less than 100 cases and those countries that began reporting cases only recently (end of March) were excluded from the sampling frame. Hence the sampling frame was finally reduced to 105 countries.

A sampling frame of 105 countries with a 10 % marginal error and 95% confidence interval yielded a total sample size of 50 countries. Hence a purposive sampling of 50 countries across the globe was carried out to have fair representation from different geographical locations.

**Fig 1:**
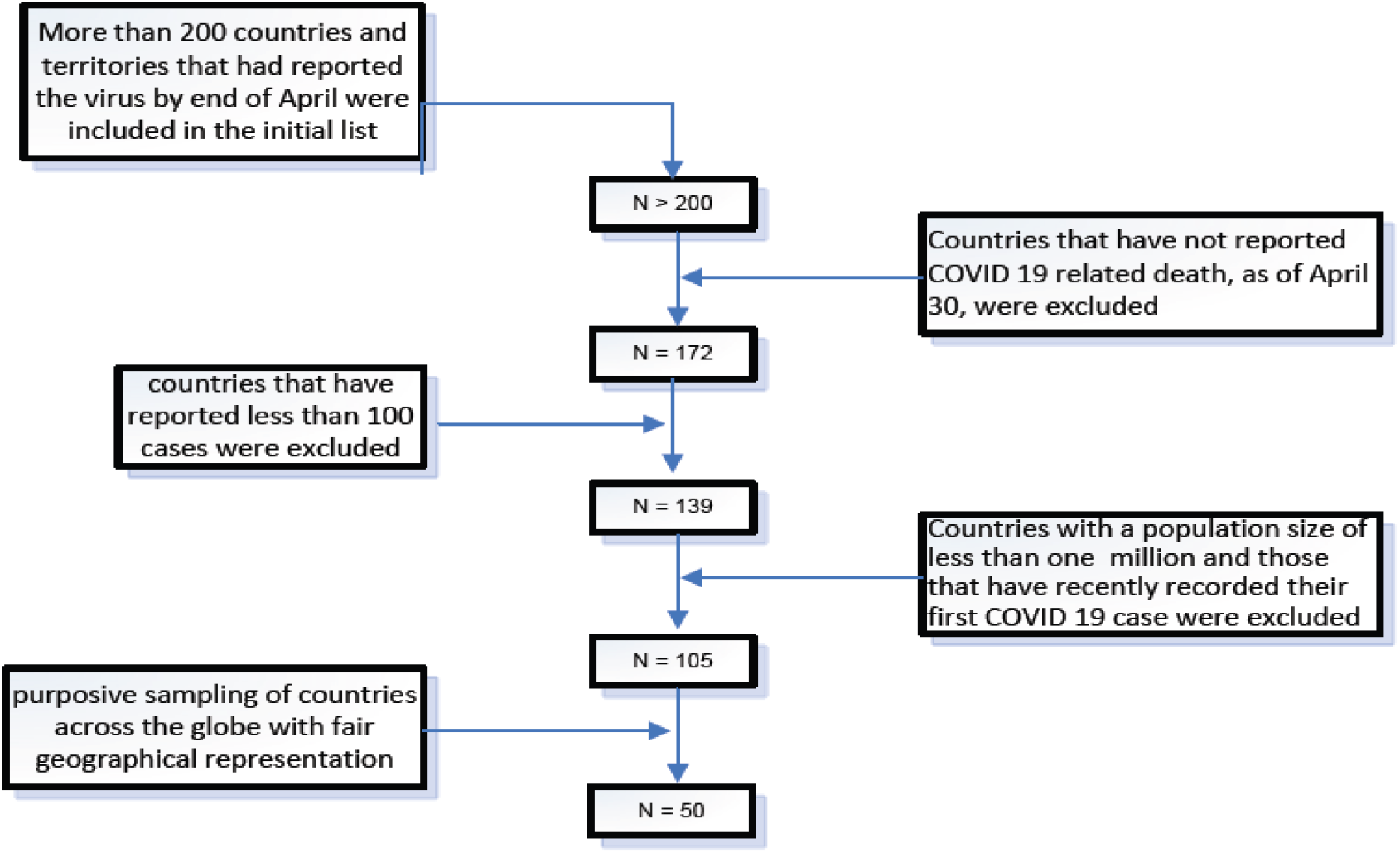
Sampling procedure.

### Data source

We used data from the COVID-19 data repository of the Johns Hopkins Center for Systems Science and Engineering (Baltimore, MD, USA) to obtain the cumulative number of cases since the diagnosis of the first patient by country. Data from the European center for disease control and prevention and the World Health Organization’s daily situational report were used to obtain the daily number of cases and organize the incidence data. [15] The 2018 World bank report was used to obtain data on the demographic distribution given by the proportion of the elderly population, and population density given by the total number of people living per square kilometer of area for each country that are included in the study. [16]

### Analysis approach

The two major variables of the study namely disease transmission and disease fatalities were assessed for the fifty countries sampled using the latest data from the above data sources.

The first variable, disease severity (fatality), was analyzed using the case fatality ratio of COVID-19, which is defined as the proportion of people with COVID-19 (that is, with a positive test result) who die as a direct or indirect consequence of their infection. [17] Despite its tendency to overestimate fatality due to undertesting and a time-lag bias the CFR seems to remain the best tool to express the fatality of the disease, even though it might underestimate this figure in the initial phase of an outbreak. In addition to the reported case fatality ratio, the pooled estimate of CFR was calculated for the sampled countries. The proportion of deaths to the total numbers of cases was meta-analyzed using the R statistical software’s metaphor package, using random-effect inverse-variance weighting to estimate the case fatality ratio. To see the potential association that CFR with other factors, a multiple linear regression model was fitted and the effect of demography (explained by the proportion of population above 65 years old) and population density was analyzed.

The second variable, disease spread, was assessed using the reproduction number of the disease across the sampled countries. In doing so the R statistical software’s R0 package was used to estimate the reproduction number of the COVID-19 using the exponential growth method given by the equation

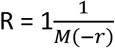

*where M is the moment generating function of the (discretized) generation time distribution, and r is the exponential growth rate. [18]*

A time frame in each country’s epidemiologic curve, where the growth of cases followed an exponential trajectory, was chosen for the analysis. In computing the reproductive number, an earlier study in Wuhan China, that has estimated the serial interval of the disease was used to compute the generation time, hence the serial interval was fitted with gamma distribution by taking the mean and standard deviation of the disease as 7.5 and 3.4 days respectively.[3] After computing the reproductive number of each country in the study, a multiple linear regression model was fitted by taking the R0 value as dependent variable, latitude and population density as an independent variable. In all the regression models a p-value of less than 0.05 was considered to declare a result as statistically significant.

## Results

### Disease transmission Analysis

To assess the disease spread, the reproductive number of COVID-19 was computed in all the sampled countries. The average reproductive number of the sampled countries was 2.1 [95% CI 2.018008 - 2.164926]. To see the impact of proximity to the equator (explained by latitude) and population density on the disease spread, a multiple linear regression model was fitted by having the R0 of the sampled countries as a dependent variable with latitude and population density as an independent variable. The regression output revealed that there is no statistically significant association between the disease spread, explained by the reproductive number, latitude and population density.

**Table 1:** Regression output for R0 and the two covariates.

| Variable | Estimate | P value | 95% CI |
| --- | --- | --- | --- |
| Latitude (proximity to the equator) | 0.004 | 0.302 | -0.00357 - 0.01127 |
| Population density | 0.00005 | 0.445 | -0.000072 - 0.00016 |

**Fig 2:**
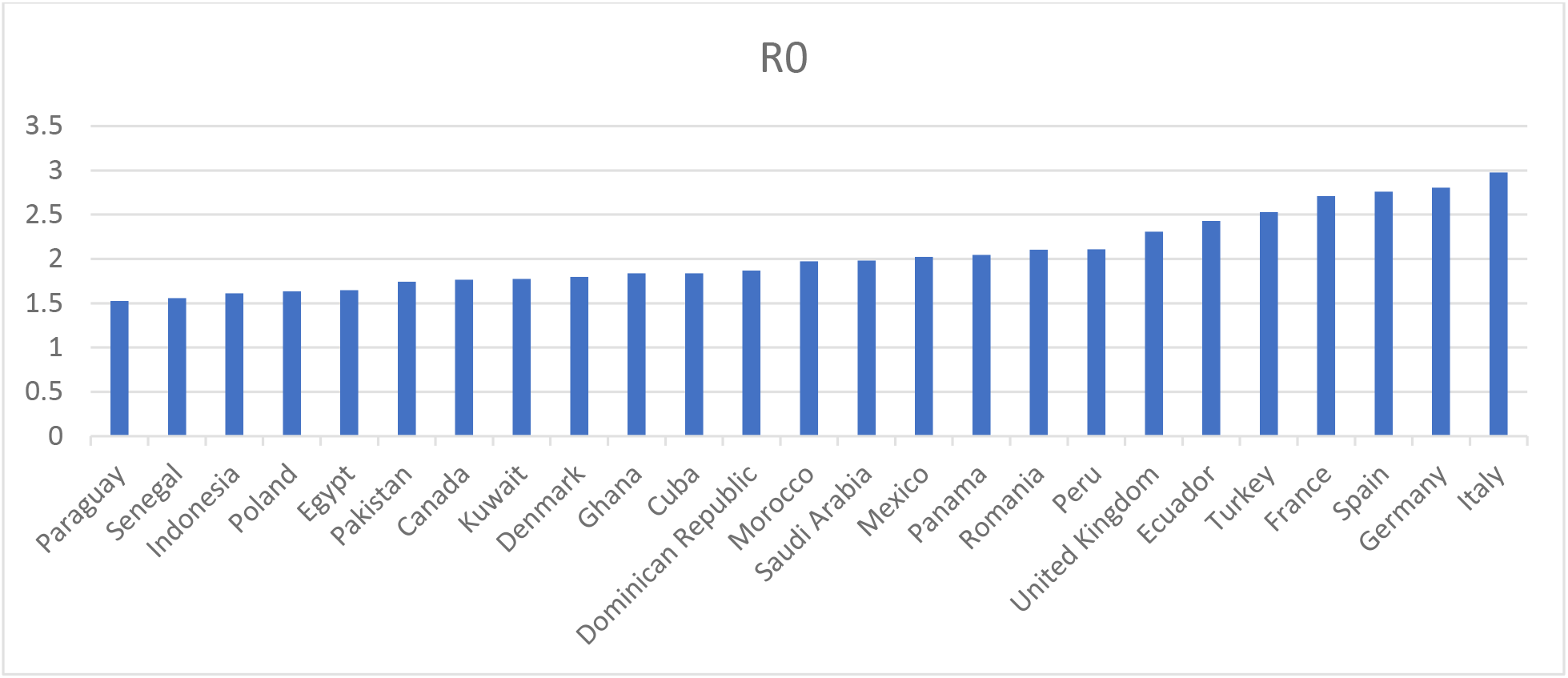
Country R0 estimates.

**Fig 3:**
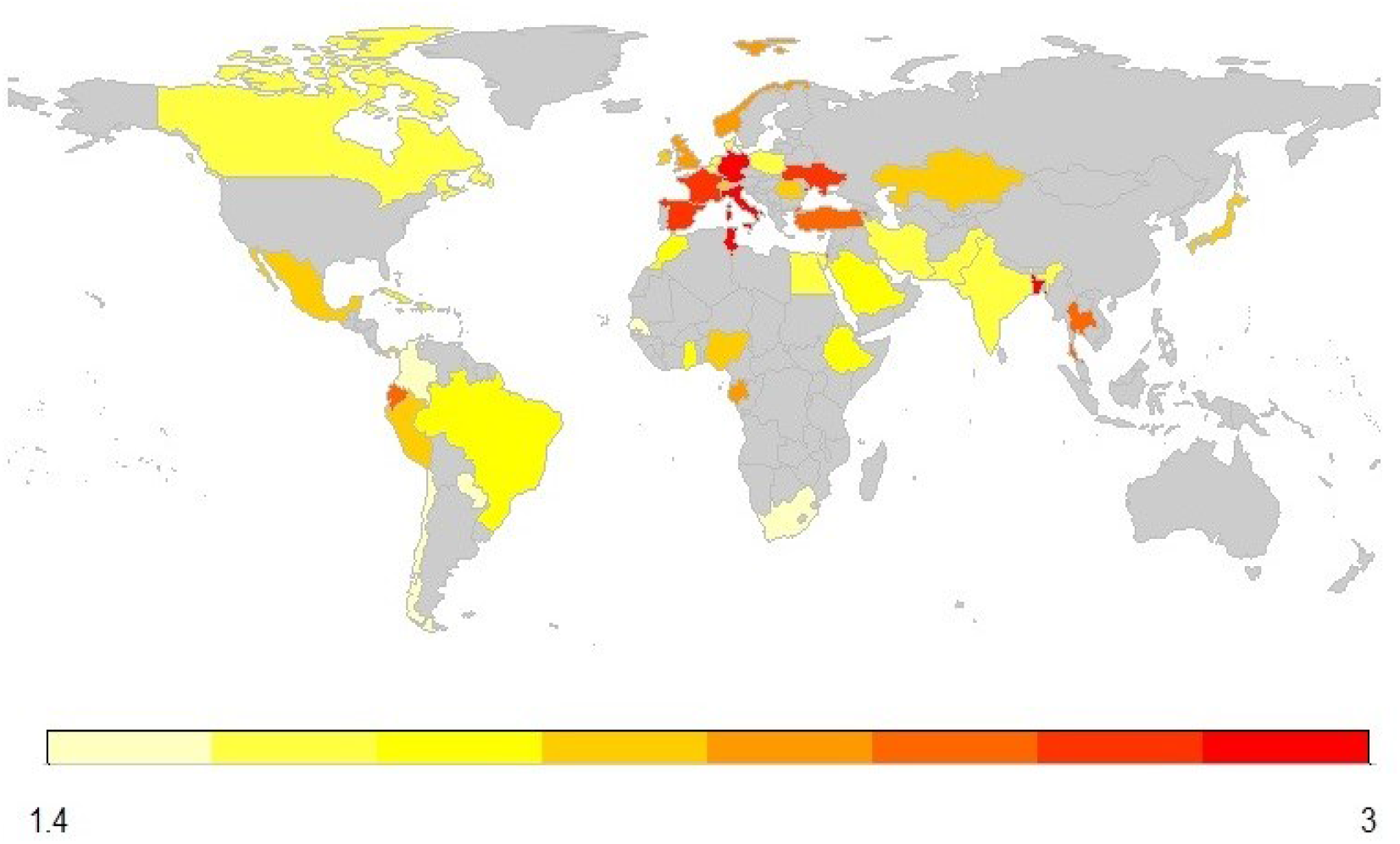
Map shows the sampled countries based on their R0 value.

**Fig 4:**
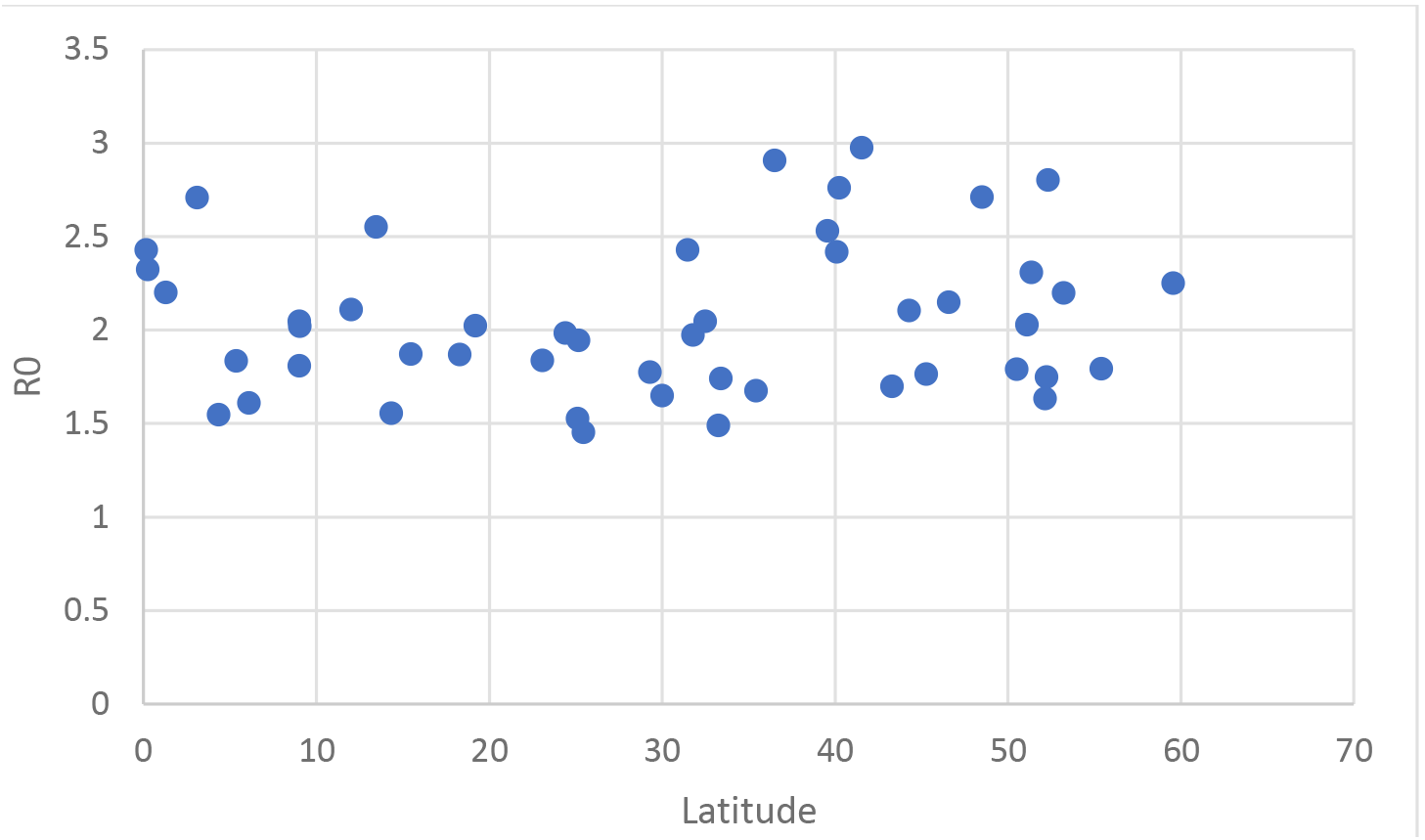
R0 estimates plotted against country latitude.

### Case fatality analysis

The random effect meta-analysis resulted in a pooled case fatality ratio of 3.32% [95% CI: 2.85, −3.86;]. The pooled overall estimate of the case fatality (weighted average) among the sampled countries is lower than the arithmetic mean of CFR (4.62 and 3.86 for the arithmetic mean and pooled estimate respectively) and this can be explained by the high heterogeneity between the studied countries. As it is illustrated on figure), the 50 countries covered in this study has exhibited a noticeable difference in the case fatality ratio, ranging from 15% in Belgium to less than 1% in some African countries.

**Table 2:** Regression output for CFR and the explanatory variables (demography and population density)

| Variable | estimate | P-value | 95% CI |
| --- | --- | --- | --- |
| **Demography (defined by the proportion of elderly population)** | 0.3231 | 0.0000377*** | 0.1803 - 0.4659 |
| **Population density** | -0.00055 | 0.1949 | -0.0013 - 0.00029 |

\*statistically significant association

The multiple linear regression model fitted between CFR and the three major covariates showed that, the demographic distribution of the sampled countries is strongly associated with the case fatality ratio. Countries that have a very high proportion of elderly population (proportion of population above 65 years of age) were found to have a higher case fatality rate. An increase in the elderly population proportion by 1% was associated with an increase in CFR by 0.32.

**Figure 5:**
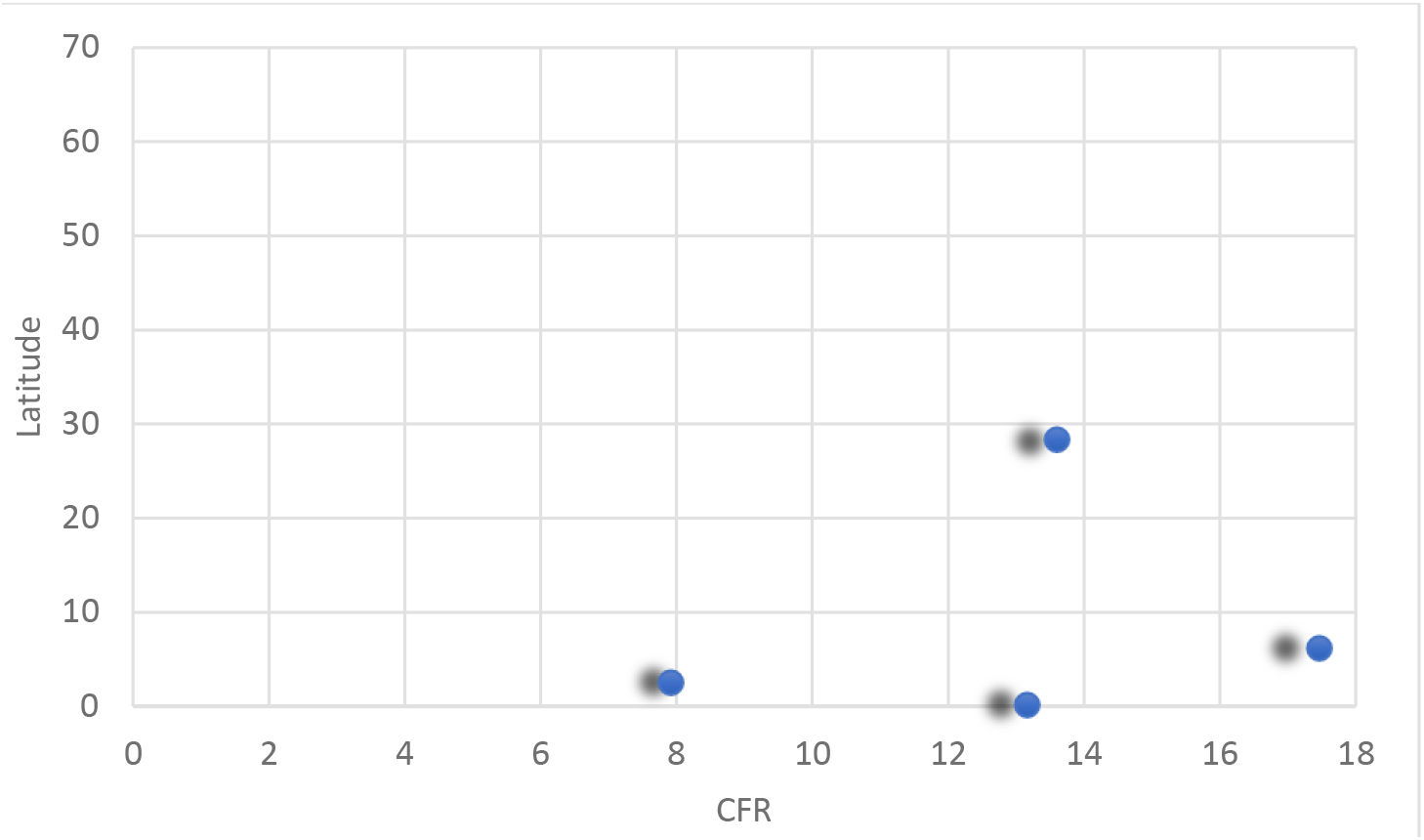
case fatality ratio by latitude.

**Fig 6:**
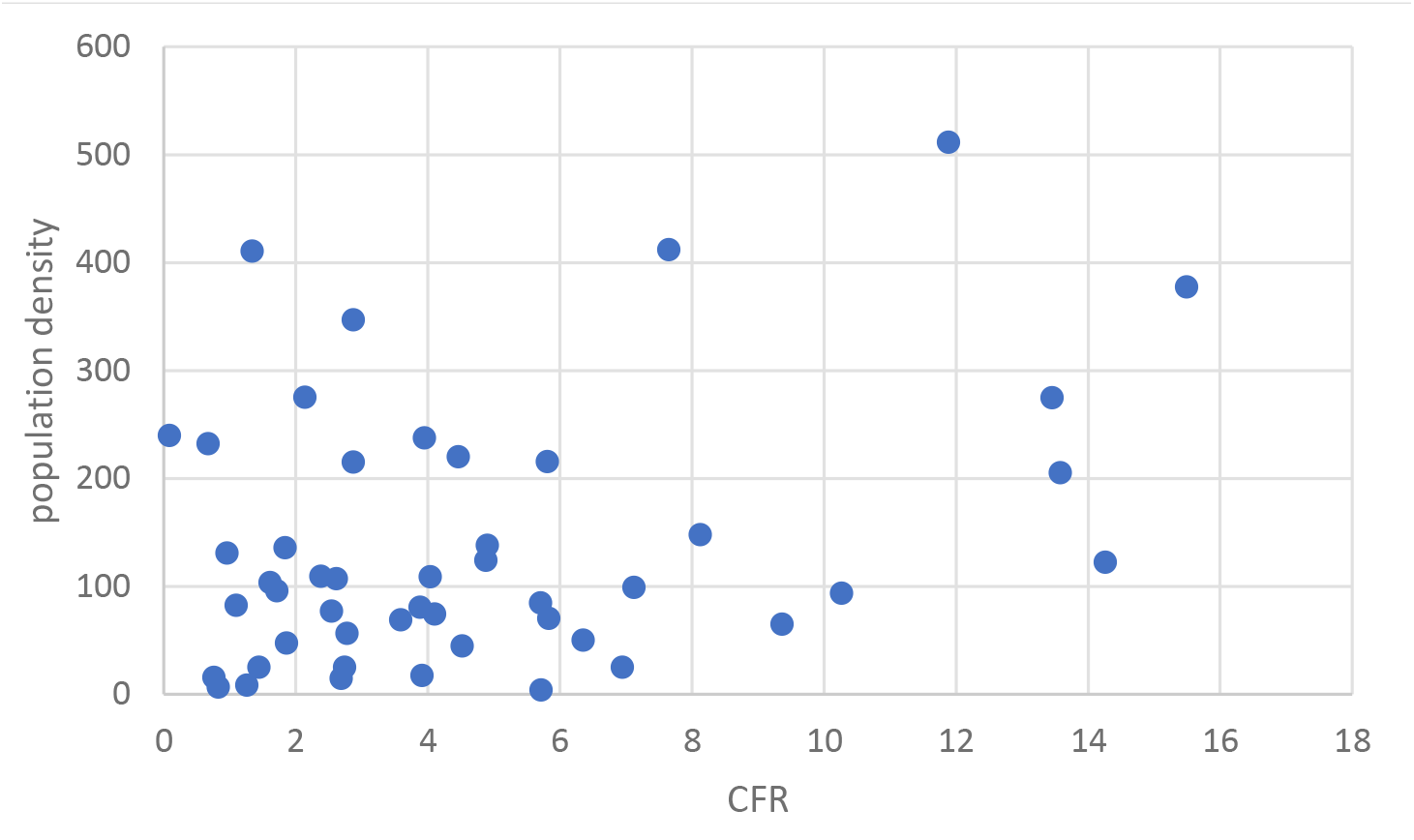
Country case fatality ratio plotted against population density.

**Fig 7:**
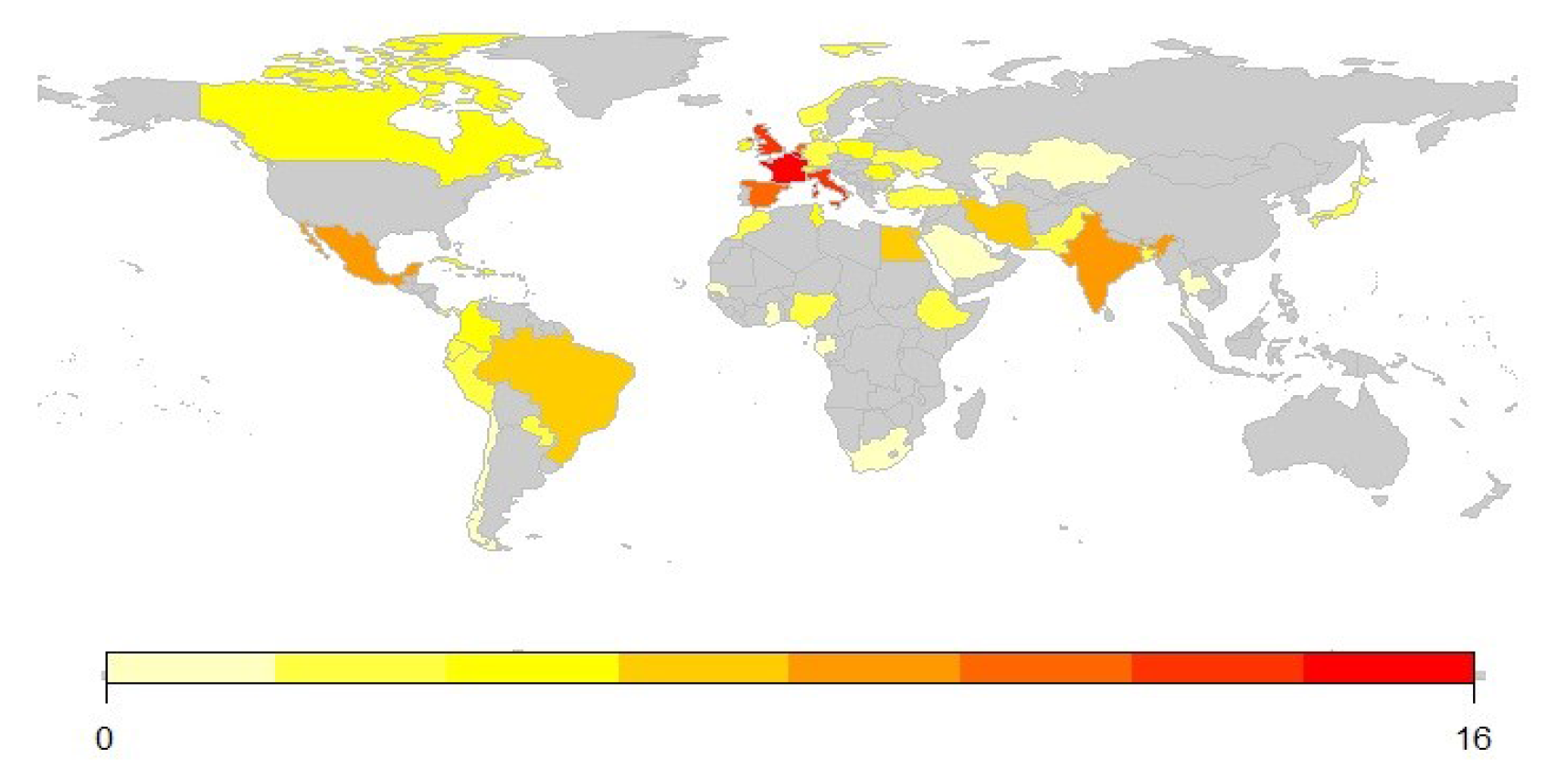
Map shows the sampled countries based on their R0 value (disease spread)

**Figure 8:**
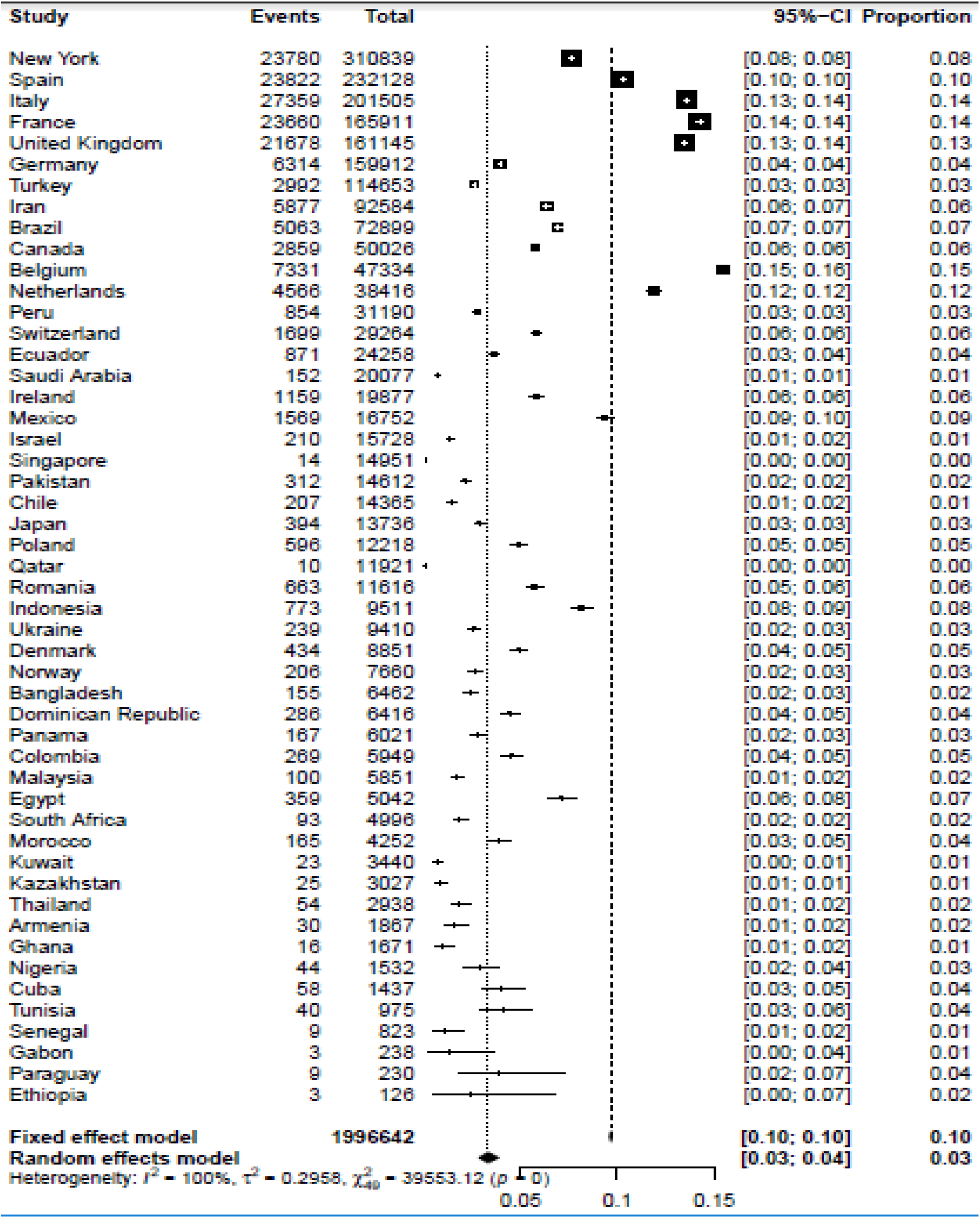
Country-level case fatality ratios.

## Discussion

Several studies that have assessed the epidemiology of COVID-19 principally focused on variables such as pollution, underlying comorbidities in infected individual, and other socioenvironmental factors to identify potential risk factors correlated with disease transmission and mortality. [19 – 25] Although these previous works have examined the effects of these variables on COVID-19 spread and fatality, most of the studies looked at effects of independently and did not control for the potentially confounding interactions between variables. Parameters of the COVID-19 pandemic that we have analyzed herein are important for predicting the spread and managing the ongoing outbreak. Particularly, cross-country comparisons of CFR and RR as important indicators of disease spread and severity are vital for setting priority for public health response and monitoring health system capacity and performance. Nevertheless, the magnitude of undetected cases or the length of delay in case reporting can significantly affect CFR and RR and can confound the estimations.

In this study, we sought to investigate COVID-19 spread and mortality worldwide through a more broader framework that accounts for effects of age distribution, population density, and latitude variables to show important correlations for the epidemiological parameters of the 50 countries or territories analyzed. By conducting sequential multivariate analyses of these variables our study aims to more comprehensively characterize the epidemiology of SARS-CoV-2 and thereby identify particular noticeable risk factors and vulnerable population groups.

A number of studies have established that in the Northern Hemisphere, influenza is more widely spread during the winter seasons, and that influenza virus transmission and virulence depend also on meteorological conditions such as temperature and relative humidity. A similar behavior has been observed for other SARS coronaviruses that belong to the same Coronaviridae family of SARS-CoV-2. [26 – 27]

Our study included data from a range of countries and climatic regions. Our regression model analysis that accounted for age, population density and latitude showed that age distribution remains an important driver shaping the current distribution of COVID-19 cases. The relative frequency of people ≥ 65 years old was positively correlated with the cumulative numbers of COVID-19 cases as well as case fatality ratio in each country. Correlation with proportion of populations over 65 years old is concordant with the previous findings relationship between case fatality ratio and patient age. [9] In other previous studies environmental factors that influence the relationship between temperature and mortality, such as latitude, humidity, and sociodemographic factors have been reported. [28 – 30]

We found no significant association for the parameters analyzed with population density. Previously spotted correlation of population density with virus spread and mortality requires further investigation before concluding that there is a correlation. We also examined possible association of incidence and mortality with country centroid latitude. Although human mobility and host susceptibility were main drivers in the spread of COVID-19, the potential role of latitude as a factor of COVID-19 burden was not further supported by our finding regarding latitude which did not suggest any protective effect of tropical climates. However, other studies analysis suggests that hemisphere might influence mortality (the latter seems to be higher in the Northern Hemisphere, p = 0.036) and thus does not contradict the idea that outdoor temperature could impact chances for patient recovery. [31] Other studies have also reported that temperature and humidity are associated with a higher risk of COVID-19. [32] Therefore, although humidity was not considered in our research because of the lack of sufficient information on this variable, precipitation seems to be an important factor that must be further examined.

## Limitations

Our analysis only included countries with sufficient data, and relied heavily on publicly available sources, which may not always be consistent with the actual value of various statistics for each countries. Many countries were excluded from analysis for lack of adequate Covid-19 cases or lack of Covid-19 deaths. This limitation is unavoidable as waiting for sufficient data may deter timely generation of evidence for appropriate public health response. This study is also limited by the fact the COVID-19 pandemic is at different stages in different countries while still spreading across the globe. The number of cases and mortality statistics we used only provide snapshots of the current state for the countries and will not be representative of the cumulative COVID-19 cases and mortality numbers for the counties. Confounding effects are also possible, given that it is impossible to fully disentangle socioeconomic and demographic variables from each other. Specifically, our study does not take into account variables relevant to local factors associated with community spread or containment measures implemented against the epidemic in individual countries.

## Conclusion

This study assessed the independent effects of latitude, age distribution and population density on the transmission rate of cases and mortality of COVID-19 worldwide. In multivariate regression analyses that controlled for country demographics, latitude and population density, countries with higher proportion of elderly population were associated with increased COVID-19 cases and deaths. Our findings showed no correlation between a country’s population density and the number of COVID-19 cases or mortality, and no correlation between latitude and mortality or rate of spread of COVID-19.

This analysis provides important information that can inform the decisions of local and global health authorities. Particularly, as our study confirms that death and severity of COVID-19 are associated with age, in countries with the biggest outbreaks, strategies must be employed to ensure that high-risk groups, such as old people received adequate protection from COVID-19.

## Data Availability

Available upon request

## References

1. Novel Coronavirus (COVID-19) Cases, provided by the Johns Hopkins Center for Systems Science and Engineering. Available: https://coronavirus.jhu.edu/map.html. Accessed on May 10, 2020.

2. Statement on the second meeting of the International Health Regulations (2005) Emergency Committee regarding the outbreak of novel coronavirus (2019-nCoV). Available: https://www.who.int/news-room/detail/30-01-2020-statement-on-the-second-meeting-of-the-international-health-regulations-(2005)-emergency-committee-regarding-the-outbreak-of-novel-coronavirus-(2019-ncov). Accessed on May 10, 2020

3. Li, Q., Guan, X., Wu, P., Wang, X., Zhou, L., Tong, Y., Ren, R., Leung, K. S. M., Lau, E. H. Y., Wong, J. Y., Xing, X., Xiang, N., Wu, Y., Li, C., Chen, Q., Li, D., Liu, T., Zhao, J., Liu, M.,... Feng, Z. (2020). Early transmission dynamics in Wuhan, China, of novel coronavirus-infected pneumonia. In New England Journal of Medicine (Vol. 382, Issue 13, pp. 1199–1207). Massachussetts Medical Society. https://doi.org/10.1056/NEJMoa2001316

4. Lu R, Zhao X, Li J, et al. Genomic characterization and epidemiology of 2019 novel coronavirus: implications for virus origins and receptor binding. Lancet. 2020;395(10224):565–574. doi:10.1016/S0140-6736(20)30251-8

5. Althaus C. Estimating case Fatality Ratio of COVID-19 from Observed Cases outside China. [(accessed on 16 February 2020)]; Available online: https://github.com/calthaus/ncov-cfr. Accessed on…

6. Wu, J.T., Leung, K., Bushman, M. et al. Estimating clinical severity of COVID-19 from the transmission dynamics in Wuhan, China. Nat Med 26, 506–510 (2020). https://doi.org/10.1038/s41591-020-0822-7

7. N. M. Ferguson, et al., Impact of non-pharmaceutical interventions (NPIs) to reduce COVID-19 mortality and healthcare demand (2020). Report of Imperial College COVID19 Response Team, March 16,2020.

8. Ficetola G.F. and Rubolini D. (2020) Climate affects global patterns of Covid-19 early outbreak dynamics. medRxiv preprint doi: https://doi.org/10.1101/2020.03.23.20040501

9. Onder, Graziano, Giovanni Rezza, and Silvio Brusaferro. 2020. “Case-Fatality Rate and Characteristics of Patients Dying in Relation to COVID-19 in Italy.” JAMA: The Journal of the American Medical Association, March. https://doi.org/10.1001/jama.2020.4683.

10. Braiman, Mark, Latitude Dependence of the COVID-19 Mortality Rate—A Possible Relationship to Vitamin D Deficiency? (March 26, 2020). Available at SSRN: https://ssrn.com/abstract=3561958 or http://dx.doi.org/10.2139/ssrn.3561958

11. Triplett, Michael. 2020. “Evidence That Higher Temperatures Are Associated with Lower Incidence of COVID-19 in Pandemic State, Cumulative Cases Reported up to March 27, 2020.” medRxiv, April, 2020.04.02.20051524.

12. United Nations, World Population Prospects 2019. https://population.un.org/wpp/DataQuery/. Accessed 13 March 2020.

13. Salmon, Why are Korea’s COVID-19 death rates so low? Asia Times, 11 March 2020. https://asiatimes.com/2020/03/why-are-koreas-covid-19-death-rates-so-low/. Accessed 13 March 2020.

14. Martinez-Alvarez M, Jarde A, Usuf E, et al. COVID-19 pandemic in west Africa. The Lancet. Global Health. 2020 May;8(5):e631-e632. DOI: 10.1016/S2214-109X(20)30123-6.

15. COVID-19 Global overview. Data on Geographic Distribution of COVID-19. European Centre for Disease Prevention and Control (ECDC). Available: https://qap.ecdc.europa.eu/public/extensions/COVID-19/COVID-19.html Accessed on April 30, 2020

16. The World Bank Open Data. Available: https://data.worldbank.org/indicator/sp.pop.totl Accessed on April 30, 2020

17. World Health Organization. The First Few X cases and contacts (FFX) investigation protocol for coronavirus disease 2019 (COVID-19). WHO/2019-nCoV/FFXprotocol/2020.2

18. Wallinga, J., & Lipsitch, M. (2007). How generation intervals shape the relationship between growth rates and reproductive numbers. Proceedings of the Royal Society B: Biological Sciences, 274(1609), 599–604. https://doi.org/10.1098/rspb.2006.3754

19. Garg S KL, Whitaker M, et al. Hospitalization Rates and Characteristics of Patients Hospitalized with Laboratory-Confirmed Coronavirus Disease 2019 — COVID-NET, 14 States, March 1–30, 2020. MMWR Morb Mortal Wkly Rep. 2020.

20. Zhang J, Litvinova M, Wang W, et al. Evolving epidemiology and transmission dynamics of coronavirus disease 2019 outside Hubei province, China: a descriptive and modelling study. Lancet Infect Dis. 2020.

21. Zhong BL, Luo W, Li HM, et al. Knowledge, attitudes, and practices towards COVID-19 among Chinese residents during the rapid rise period of the COVID-19 outbreak: a quick online cross-sectional survey. Int J Biol Sci. 2020;16(10):1745–1752.

22. Wu X, Nethery RC, Sabath BM, Braun D, Dominici F. Exposure to air pollution and COVID-19 mortality in the United States. medRxiv. 2020:2020.2004.2005.20054502.

23. Gardner W, States D, Bagley N. The Coronavirus and the Risks to the Elderly in Long-Term Care. J Aging Soc Policy. 2020: 1–6.

24. Jordan RE, Adab P, Cheng KK. Covid-19: risk factors for severe disease and death. BMJ. 2020;368:m1198.

25. Zhou F, Yu T, Du R, et al. Clinical course and risk factors for mortality of adult inpatients with COVID-19 in Wuhan, China: a retrospective cohort study. Lancet (London, England). 2020;395(10229):1054–1062.

26. Lowen AC, Mubareka S, Steel J, Palese P. Influenza Virus Transmission is Dependent on Relative Humidity and Temperature. PLoS Pathol, 2007 Oct; 3(10): e151.doi: https://dx.doi.org/10.1371/journal.ppat.003015115.

27. Prussin AJ, Schwake DO, Lin K, Gallagher DL, Buttling L, Marr LC. Survival of the Enveloped Virus Phi6 in Droplets as a Function of Relative Humidity, Absolute Humidity, and Temperature. Appl Environ Microbiol, 2018 May 31;84(12). pii: e00551–18. https://dx.doi.org/10.1128/AEM.00551-18

28. Bao, J., Wang, Z., Yu, C., Li, X., 2016. The influence of temperature on mortality and its Lag effect: a study in four Chinese cities with different latitudes. BMC Public Health 16 (1), 375–383. https://doi.org/10.1186/s12889-016-3031-z.

29. Jaakkola, K., Saukkoriipi, A., Jokelainen, J., Juvonen, R., Kauppila, J., Vainio, O., Ikäheimo, T.M., 2014. Decline in temperature and humidity increases the occurrence of influenza in cold climate. Environ. Health 13 (1), 22.

30. Kudo, E., Song, E., Yockey, L.J., Rakib, T., Wong, P.W., Homer, R.J., Iwasaki, A., 2019. Low ambient humidity impairs barrier function and innate resistance against influenza infection. Proc. Natl. Acad. Sci. 116 (22), 10905–10910. https://doi.org/10.1073/pnas.1902840116

31. Shagam L. Untangling factors associated with country-specific COVID-19 incidence, mortality and case fatality rates during the first quarter of 2020. medRxiv; 2020. DOI: 10.1101/2020.04.22.20075580.

32. J. Wang, K. Tang, K. Feng, W. Lv, High Temperature and High Humidity Reduce the Transmission of COVID-19 SSRN, (2020).

